# Leading causes of mortality and prescription drug coverage in Canada and New Zealand

**DOI:** 10.1101/2020.02.17.20024232

**Authors:** Nigel S B Rawson

## Abstract

**Background:** Canada may soon see the introduction of a national pharmaceutical insurance system. New Zealand has a government-funded healthcare system used by all residents that operates within a tight cost-containment budget.

**Objective:** To compare the main mortality causes in Canada and New Zealand and examine listings in current Canadian provincial public drug plans and the New Zealand national drug formulary.

**Methods:** Age-standardized mortality rates from 1985 to 2014 and data on hospital discharges and average length of stay in hospital for Canada and New Zealand were obtained from the Organization for Economic Cooperation and Development’s website. Information on insured medications was obtained from Canadian provincial drug plan lists and the New Zealand Pharmaceutical Schedule current in mid-2019.

**Results:** Mortality rates for acute myocardial infarction, ischemic heart disease and cerebrovascular disease were higher, on average over the 30-year observation period, in New Zealand, but rates for malignancies and respiratory disorders were similar in the two countries. Reimbursement listing rates for drugs for cancer and some cardiovascular indications were lower in New Zealand than in Canada.

**Conclusions:** New Zealand’s drug coverage system has contained costs, but it restricts or denies access to new innovative medicines with the potential to improve patients’ lives. Although a New Zealand-style national pharmacare scheme in Canada would offer the opportunity to restrain drug expenditure, it would likely fail to satisfy patients and healthcare providers and diminish health outcomes, resulting in higher costs in other healthcare sectors.

## Background

The sustainability of prescription drug insurance coverage is a perennial political issue in Canada. Provincial governments for whom healthcare consumes close to 50% of their overall budgets are concerned about new innovative medications that provide therapies for conditions not previously treatable but have high costs. Since it is easier financially and politically to attempt to contain medication expenses than hospital and healthcare provider costs, governments commonly focus on controlling prescription drug costs.

The Canadian pharmaceutical environment will soon see significant changes as part of the federal government’s focus on “affordability, accessibility and appropriate use of prescription drugs.”^1^ For example, the government has imposed sweeping changes to the regulations that govern the quasi-judicial agency that sets maximum prices for patented medicines sold in Canada.^2^ The new regulations replace countries that have higher drug prices with lower price countries in the agency’s international price comparison analysis, enforce a hard, low cost-effectiveness threshold, impose a reduction in a drug’s price if its annual sales exceed a specified amount, and require pharmaceutical companies to divulge information on confidential rebates negotiated with Canadian payers.^3^ These changes will reduce Canada’s attractiveness to pharmaceutical manufacturers as an important country in which launch their new products.^4^

In addition, interest in a national drug insurance program has intensified with the announcement in its 2019 budget that the Liberal federal government is “moving forward” on “foundational elements” of national pharmacare.^5^ With the re-election of the Liberals, the process is likely to proceed. Canada is the only country in the world with a universal public health system that covers healthcare providers, hospitalizations and laboratory services but not prescription drugs in the outpatient setting. Reimbursement for drug expenditures is available through government-funded plans and private insurance paid for by individuals or cost-shared with employers or unions. Around two-thirds of Canadians are covered by private insurance, while federal and provincial government drug plans – mainly designed to provide coverage to seniors, social assistance recipients and some special groups, or when costs are deemed to be catastrophic – offer a degree of coverage to about 25% of the population.^6,7^ However, government plans have complex systems of deductibles, copayments and premiums and, for many drugs, special or restricted access criteria that results in variation in patient eligibility, out-of-pocket expenses and coverage, which has led to significant inequalities between provinces.^8^

New Zealand has a government-funded universal healthcare system that all residents use. To provide more timely access to medical and dental benefits, about 35% of the adult population has private health insurance most of whom pay for it themselves.^9^ However, most insurers in the New Zealand market only cover the copayments on prescription medicines that are publicly funded.

New Zealand’s government created the Pharmaceutical Management Agency, known as PHARMAC, in 1993 “to make decisions on which medicines and medical devices are funded in order to get the best health outcomes from within the available funding.”^10^ PHARMAC operates on a fixed budget for pharmaceuticals so that, when assessing a new medicine for inclusion in the Pharmaceutical Schedule (the drug formulary),^11^ funding decisions are based on clinical and economic assessments that examine the need and health benefit of a new medicine and its costs and potential savings.^12^ Consideration is also given to other drugs that must be forgone and price concessions that must be obtained from manufacturers to fund the new product within the budgetary cycle. PHARMAC takes recommendations from its health technology assessment expert committee and negotiates with manufacturers to reach a provisional listing agreement. A product is added to the national Pharmaceutical Schedule only if an acceptable proposal is achieved, which can lead to “bundling” deals with manufacturers for multiple medicines.^13^ As of June 2018, over 100 medications had been recommended for inclusion in the subsidized program by the health technology assessment committee but were unfunded, some of which had been waiting for funding for more than 10 years.^14^

## Objective

The objective of this evaluation was to review mortality and hospital discharge rates in the three disease areas (malignancies and circulatory and respiratory diseases) that account for more than two-thirds of the deaths in Canada and New Zealand in the light of prescription drug coverage by Canadian provincial public drug plans and the New Zealand national system.

## Methods

Mortality data for malignancies and circulatory and respiratory diseases for Canada and New Zealand were obtained from the website of the Organization for Economic Cooperation and Development (OECD)^15^ as age-standardized death rates per 100,000 population, standardized to the total OECD population for 2010, for the 30-year period between 1985 and 2014 (latest year for which cause of mortality data are available). Mortality data represent a hard health outcome measure.

Data on hospital discharges, excluding day cases, as a rate per 100,000 population and average length of stay in hospital, calculated by dividing the number of relevant bed-days by the number of discharges during the year, were also obtained for the two countries from 2000 to 2016 (data for both countries are only available for this period) as supplementary information about softer health outcomes.

Information on medications used to treat conditions in the three disease categories covered in Canada was obtained from on-line provincial public drug plan benefit lists current in mid-2019, including any from relevant special or exceptional access product lists maintained by some provinces. Only oncology drug reimbursement information from British Columbia, Alberta, Saskatchewan, Ontario and Quebec was used in the analysis because details of covered cancer drugs are accessible in separate publicly available benefit lists or are included in the provincial formulary for these provinces, whereas the formularies of New Brunswick, Nova Scotia, Prince Edward Island, and Newfoundland and Labrador list some but not all oncology drugs, while the oncology drug formulary for Manitoba is not publicly available. The Pharmaceutical Schedule current at mid-2019 was used to identified relevant subsidized medications, including oncology products, in New Zealand.

Where a medicine was only listed in Canadian formularies, information on regulatory applications approved by the Medicines and Medical Devices Safety Authority^16^ was used to ascertain whether the product was approved in New Zealand. Similarly, if a medication was in the New Zealand Pharmaceutical Schedule but not in any Canadian provincial formulary, Health Canada’s Drug Product Database^17^ was used to determine whether it had regulatory approval in Canada.

Fewer drugs receive regulatory approval in New Zealand,^18^ which makes comparisons of the comprehensiveness of reimbursement listing challenging. For example, if there are 20 drugs in a class of which only 10 have regulatory approval in New Zealand and all are listed in the Pharmaceutical Schedule, the reimbursement listing rate is 100%. On the other hand, if all 20 drugs have regulatory approval in Canada, but provincial drug plans only cover 10, the reimbursement listing rate is 50%, although the same number of products receive reimbursement in both countries. However, drug reimbursement systems can only include medications that have regulatory approval in their country. Consequently, only reimbursement listing rates (the number of drugs with listing as a percentage of the number with regulatory approval) are reported for each country and compared using Fisher’s exact test.

## Results

Mortality rates for Canada and New Zealand show that the main causes of death in both countries are malignancies and circulatory and respiratory diseases, accounting for 65.7% of the deaths in Canada and 71.9% of the deaths in New Zealand in 2014. The age-standardized mortality rate for circulatory diseases was higher in New Zealand than Canada by 28.2% on average, ranging between 15.9% and 37.8%, over the 30-year period, whereas the rate for malignancies was only marginally higher in New Zealand by 3.6% on average, varying between −0.3% and 8.6% (Figure 1). The age-standardized mortality rate in New Zealand from respiratory diseases also exceeded that in Canada by 41.9% on average (range: −17.7% to 101.6%) between 1985 and 1998 but was only 5.6% higher on average between 1999 and 2014 (range: −9.8% to 15.7%).

**Figure 1:**
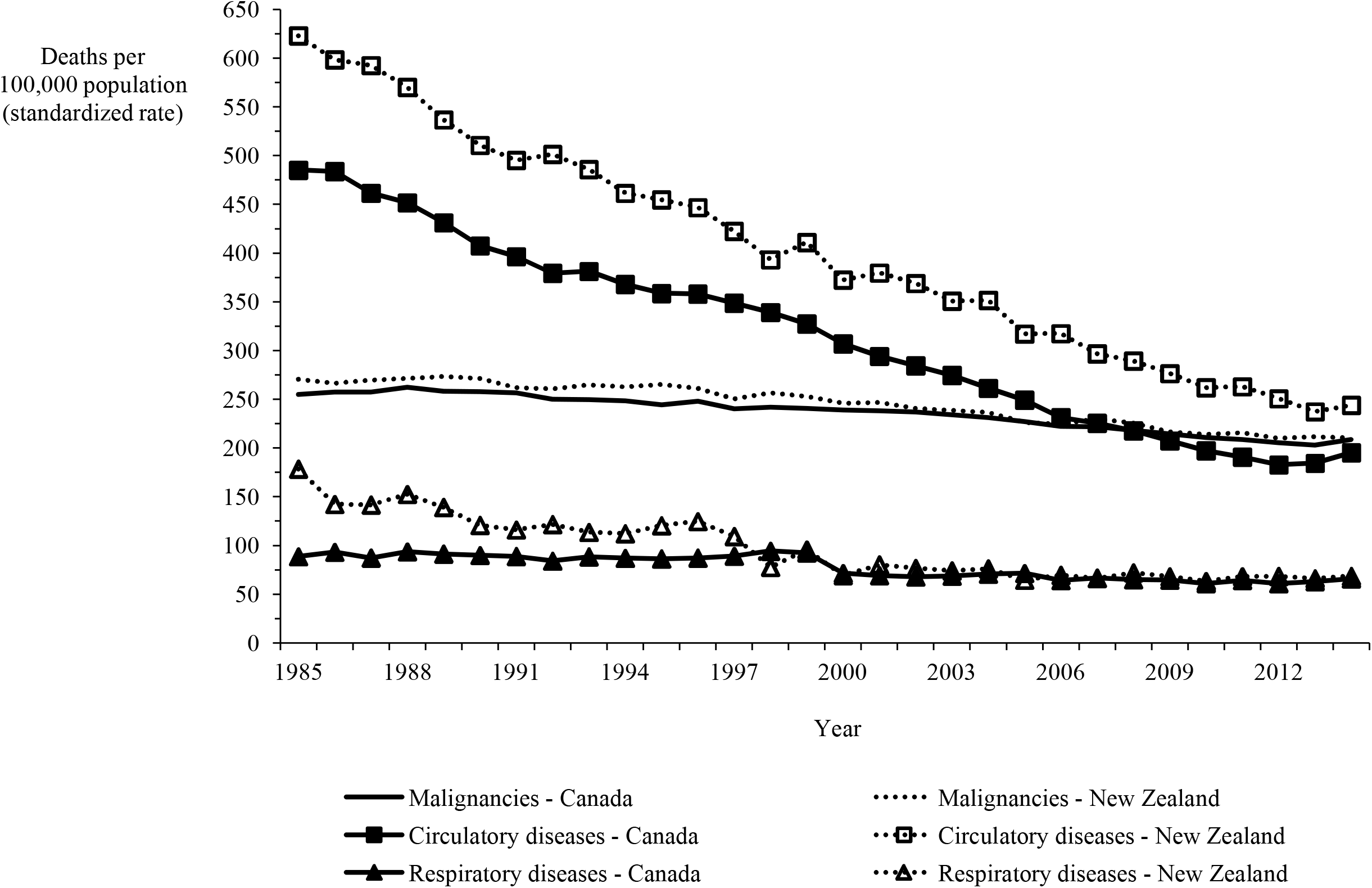
Mortality in Canada and New Zealand from malignancies and circulatory and respiratory diseases, 1985-2014.

Mortality data on ischemic heart disease and cerebrovascular disease, which account for the majority of circulatory disease deaths, demonstrate that, although the rate more than halved in both categories in both countries over the 30-year period, the rate in New Zealand exceeded the Canadian rate by 32.7% and 59.1%, on average, for ischemic heart disease and cerebrovascular disease, respectively (Figure 2). Figures 3a and 3b show mortality rates for the four major cancers: breast, colorectal, lung and prostate. The New Zealand rate was higher than the Canadian rate for breast, colorectal and prostate cancer by 9.8%, 38.7% and 31.1% on average, respectively. However, the New Zealand lung cancer rate was lower than the Canadian rate by 27.7%, on average over the 30-year period.

**Figure 2:**
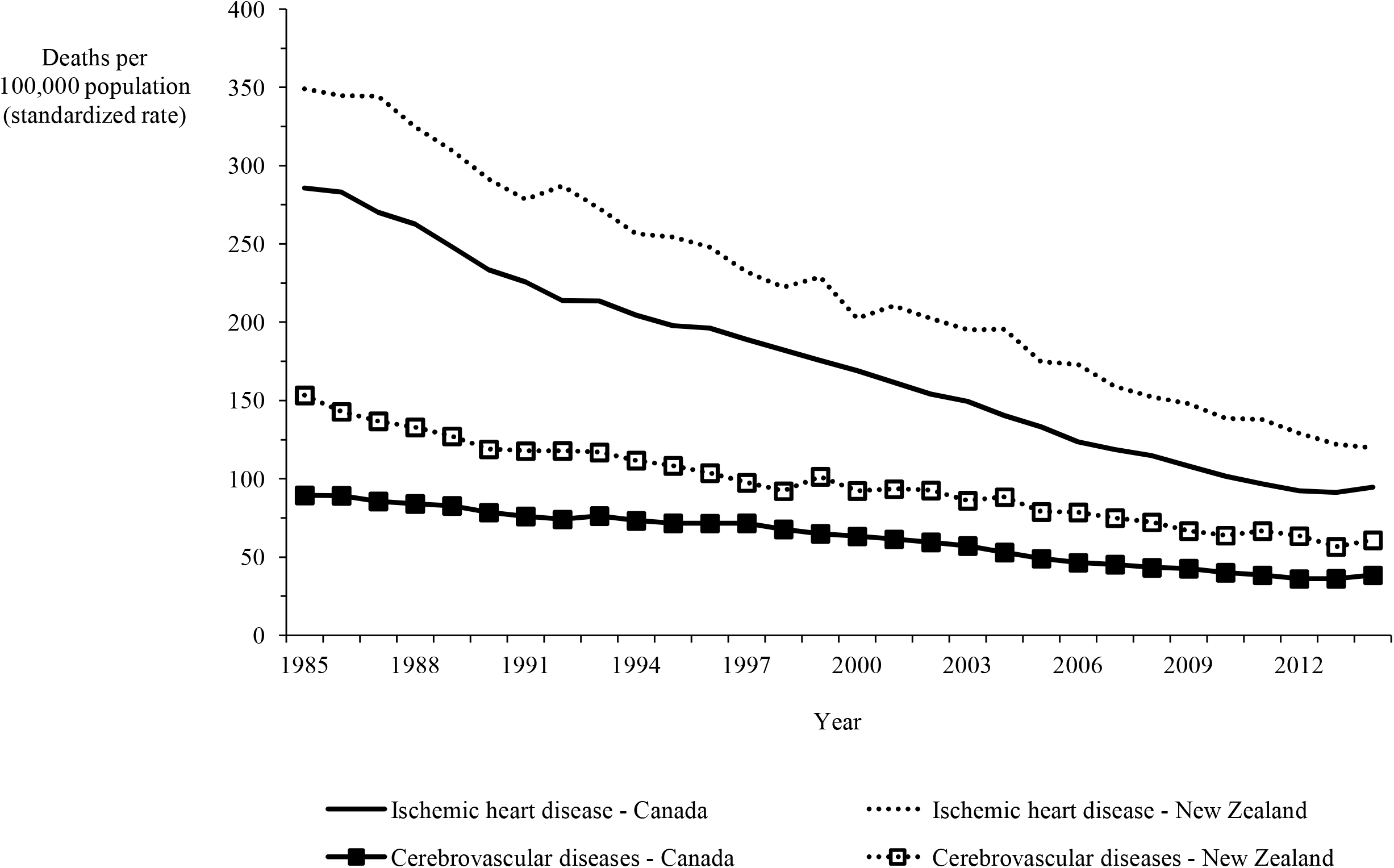
Mortality in Canada and New Zealand for ischemic heart and cerebrovascular diseases, 1985-2014.

**Figure 3a:**
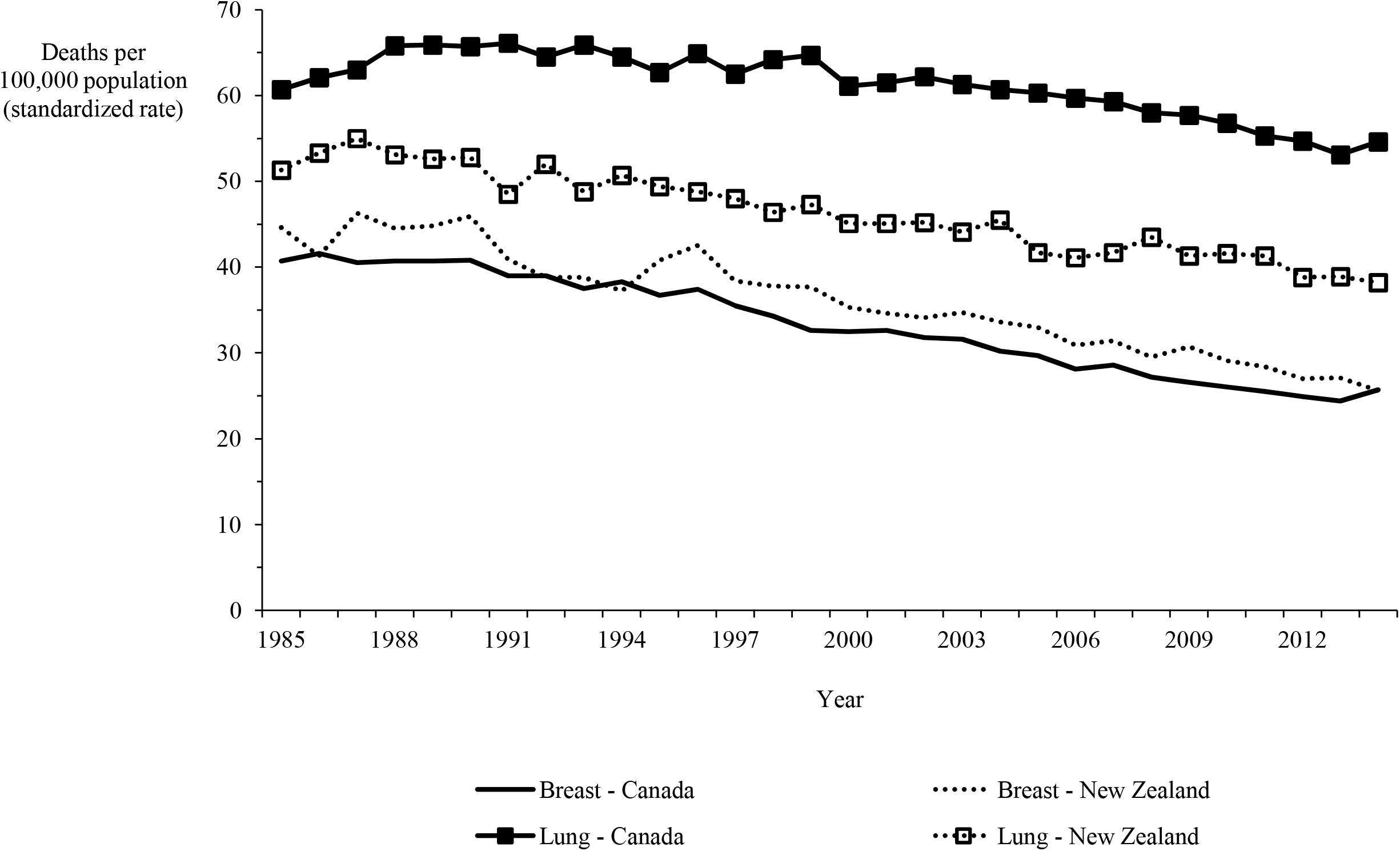
Mortality in Canada and New Zealand for breast and lung cancers, 1985-2014.

**Figure 3b:**
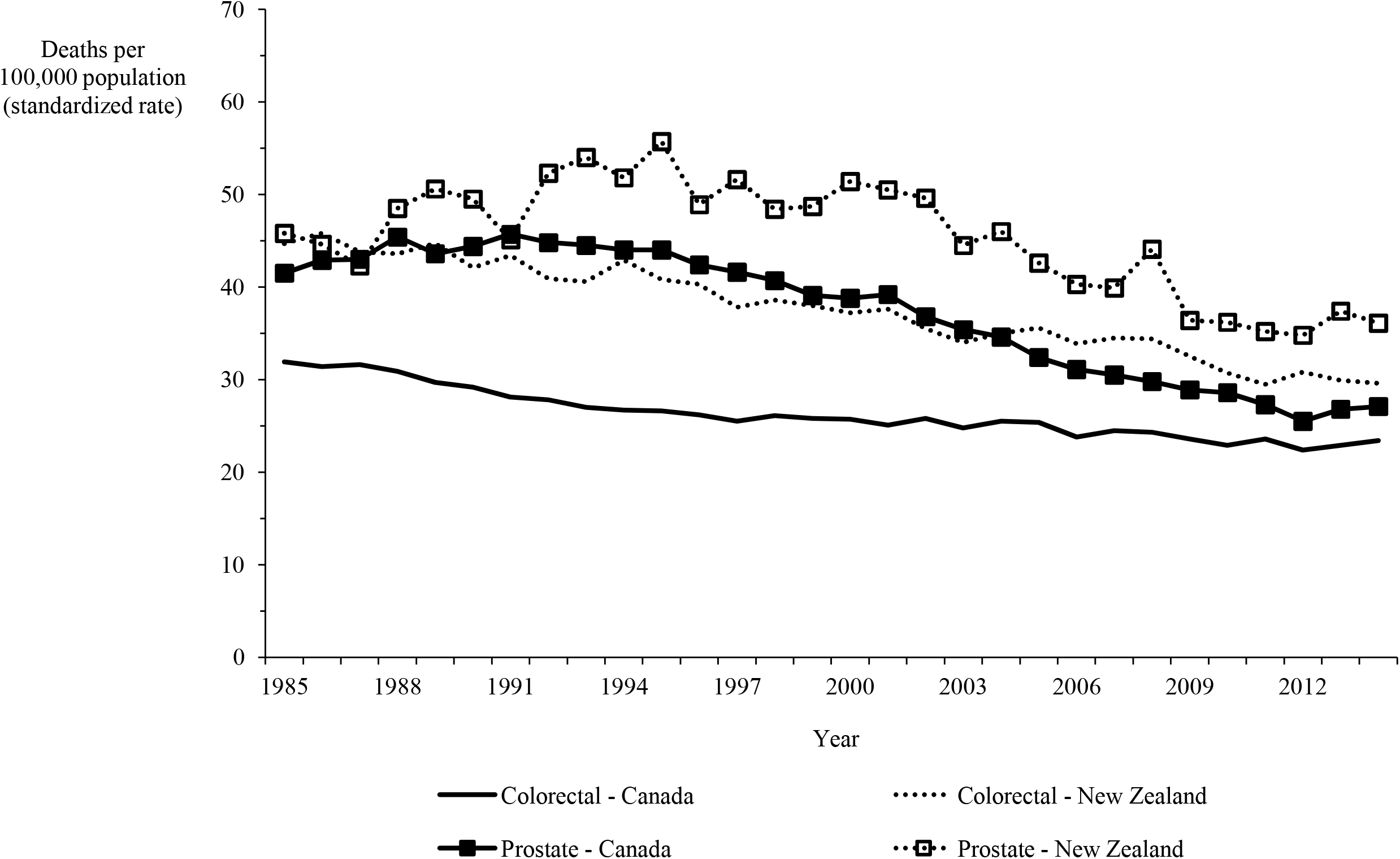
Mortality in Canada and New Zealand for colorectal and prostate cancers, 1985-2014.

Hospital discharge rates for neoplasms and circulatory diseases in New Zealand between 2000 and 2016 were consistently higher than the rates in Canada by 25% on average, while the rate for respiratory diseases was consistently higher by 67% on average (Figure 4).

**Figure 4:**
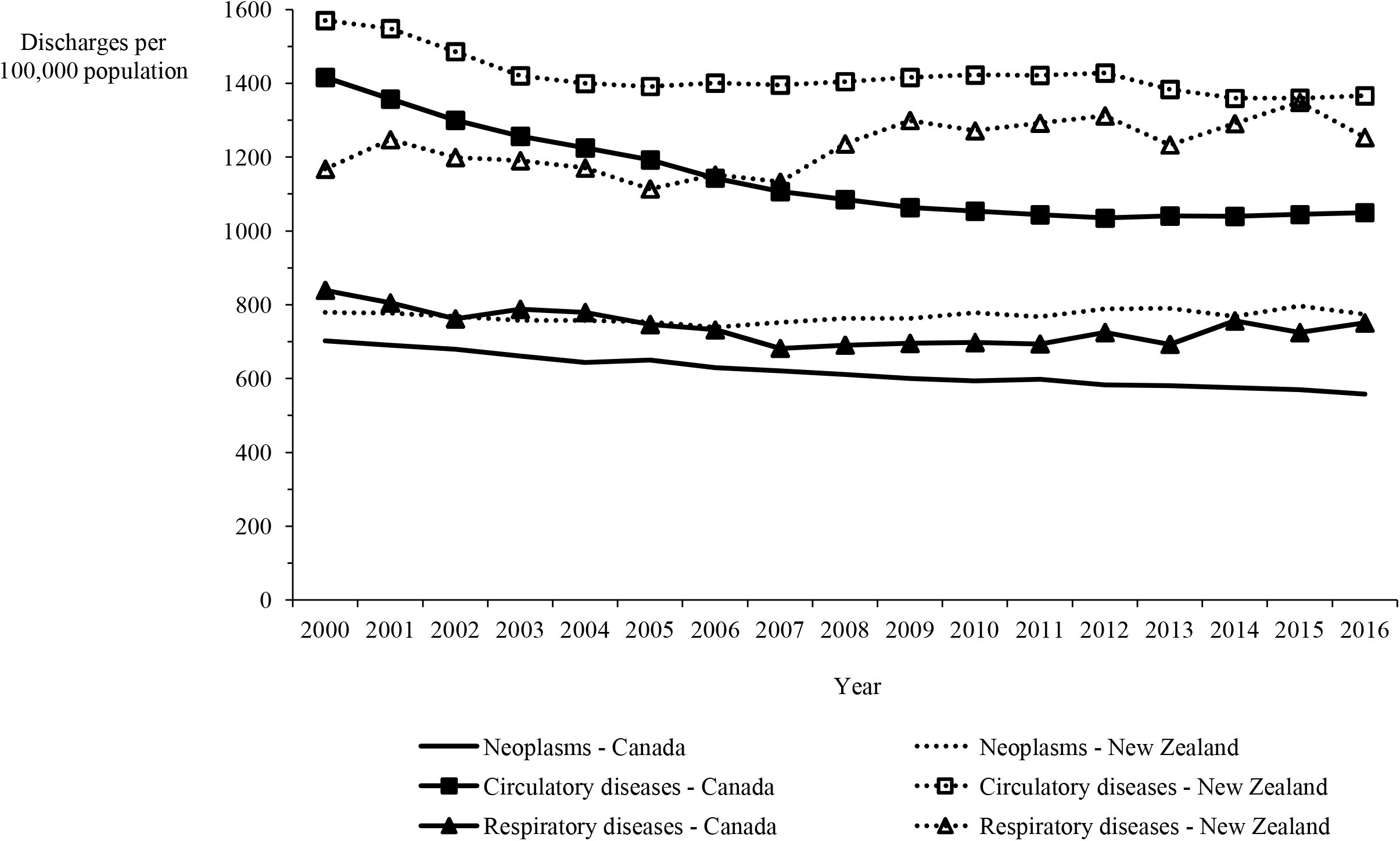
Hospital discharge rate in Canada and New Zealand for neoplasms and circulatory and respiratory diseases, 2000-2016.

Figure 5 shows the length of stay in hospital averaged over the most recent five years of data available. Although consistency exists between the two countries in the overall average length of stay and that for respiratory diseases, the New Zealand averages for disorders of the cardiovascular systems and heart failure are approximately twice those in Canada and, remarkably, the New Zealand averages for cerebrovascular diseases, ischemic heart disease other than acute myocardial infarction, and hypertensive disease are three to seven times longer than in Canada.

**Figure 5:**
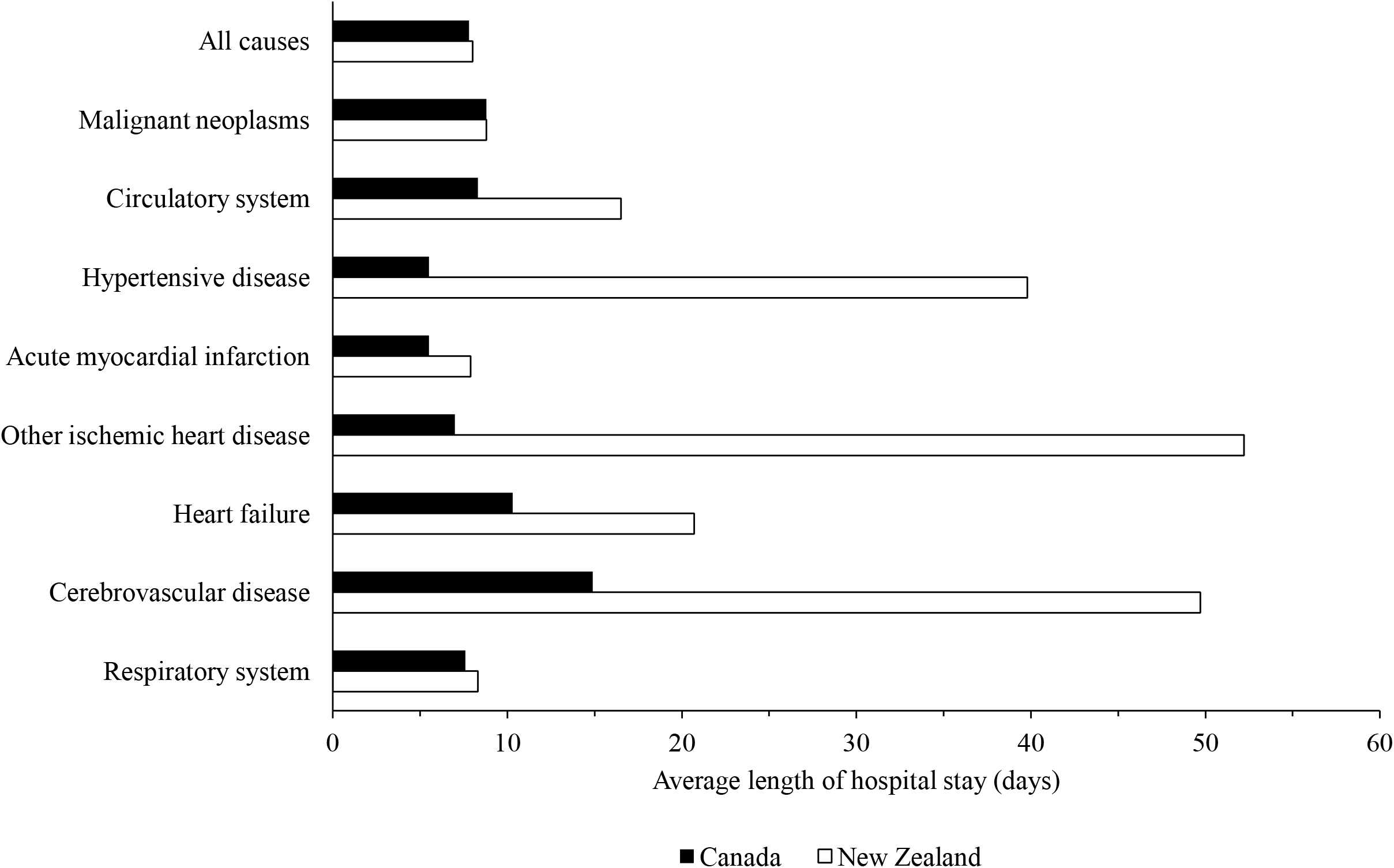
Average length of hospital stay over the most recent five years in Canada and New Zealand for all causes, malignancies, respiratory diseases and selected cardiovascular conditions.

A total of 263 medications from the three system categories (cardiovascular, anti-cancer and respiratory) were identified for which the regulatory approval rate was 85.9% in New Zealand (226 drugs) compared with 97.0% in Canada (255 drugs). Of the medicines with regulatory approval in New Zealand, 127 (56.7%) were listed in the Pharmaceutical Schedule compared with a median number of 232 drugs (91.0%) with regulatory approval in Canada that are listed in provincial drug plans (p<0.0001).

The overall reimbursement coverage rate in New Zealand was considerably lower than the median listing rate in Canadian provincial drug plans for angiotensin-converting-enzyme inhibitors (ACEIs; 60.0% v. 100.0%; p=0.007), angiotensin receptor blocking (ARB) drugs (30.8% v. 100.0%; p<0.0001), statins (71.4% v. 100.0%; p=0.19), drugs for cancers other than the four major cancer types of breast, colorectal, lung and prostate (56.2% v. 95.2%; p=0.012), and drugs used to treat multiple types of cancer (75.0% v. 96.9%; p=0.026) (Table 1). The listing rates in the two countries for other cardiovascular drugs and respiratory drugs were reasonably consistent.

**Table 1:** Summary results of the comparison of selected drugs approved and listed for benefit in New Zealand and Canada, June 2019.

| Drug group | Available | New Zealand |  |  |  | Canada |  |  |  |
| --- | --- | --- | --- | --- | --- | --- | --- | --- | --- |
|  |  | Approved for marketing No. | % | Listed for benefit in Pharmaceutical Schedule No. | % of Approved | Approved for marketing No. | % | Median No. (Range) | % of Approved (Range) |
| Cardiovascular drugs |  |  |  |  |  |  |  |  |  |
| ACEIs* | 16 | 15 | 93.8 | 9 | 60.0 | 16 | 100.0 | 16 (15-16) | 100.0 (93.8-100.0) |
| ARBs* | 16 | 13 | 81.3 | 4 | 30.8 | 16 | 100.0 | 16 (13-16) | 100.0 (81.3-100.0) |
| β-blockers* | 14 | 13 | 92.9 | 11 | 84.6 | 13 | 92.9 | 12 (10-13) | 92.3 (76.9-100.0) |
| Calcium channel blockers | 7 | 7 | 100.0 | 6 | 85.7 | 6 | 85.7 | 6 (5-6) | 100.0 (83.3-100.0) |
| Diuretics | 16 | 14 | 87.5 | 12 | 85.7 | 13 | 81.2 | 12 (9-13) | 92.3 (69.2-100.0) |
| Statins | 9 | 7 | 77.8 | 5 | 71.4 | 8 | 88.9 | 8 (7-8) | 100.0 (87.5-100.0) |
| Other anti-cholesterol | 10 | 7 | 70.0 | 7 | 100.0 | 9 | 90.0 | 7 (5-9) | 77.8 (55.6-100.0) |
| Anti-cancer drugs |  |  |  |  |  |  |  |  |  |
| Breast | 14 | 13 | 92.9 | 9 | 69.2 | 14 | 100.0 | 13 (13-13) | 92.9 (92.9-92.9) |
| Colorectal | 5 | 3 | 60.0 | 3 | 100.0 | 5 | 100.0 | 5 (5-5) | 100.0 (100.0-100.0) |
| Leukemia | 47 | 36 | 76.6 | 25 | 69.4 | 47 | 100.0 | 37 (34-41) | 78.7 (72.3-87.2) |
| Lung | 8 | 7 | 87.5 | 3 | 42.9 | 8 | 100.0 | 7 (5-8) | 87.5 (62.5-100.0) |
| Prostate | 10 | 8 | 80.0 | 4 | 50.0 | 10 | 100.0 | 9 (7-10) | 90.0 (70.0-100.0) |
| Multiple cancers | 33 | 32 | 97.0 | 24 | 75.0 | 32 | 97.0 | 31 (30-32) | 96.9 (93.8-100.0) |
| Other cancers | 21 | 16 | 76.2 | 9 | 56.2 | 21 | 100.0 | 20 (18-21) | 95.2 (85.7-100.0) |
| Respiratory drugs |  |  |  |  |  |  |  |  |  |
| LABAs† | 11 | 10 | 90.9 | 9 | 90.0 | 11 | 100.0 | 11 (10-11) | 100.0 (90.9-100.0) |
| SABAs† | 3 | 3 | 100.0 | 3 | 100.0 | 3 | 100.0 | 3 (2-3) | 100.0 (66.7-100.0) |
| Anticholinergics | 5 | 5 | 100.0 | 4 | 80.0 | 5 | 100.0 | 5 (5-5) | 100.0 (100.0-100.0) |
| Steroids | 9 | 9 | 100.0 | 8 | 88.9 | 9 | 100.0 | 9 (6-9) | 100.0 (66.7-100.0) |
| Other COPD drugs | 9 | 8 | 88.9 | 6 | 75.0 | 9 | 100.0 | 6 (3-7) | 66.7 (33.3-77.8) |

ACEI: Angiotensin-converting-enzyme inhibitor; ARB: Angiotensin receptor blocker; COPD: Chronic obstructive pulmonary disease; LABA: Long-acting beta-agonist; SABA: Short-acting beta-agonist
\* Including in combination with diuretics; † Including combinations

## Discussion

Some Canadian health policy analysts have suggested that a national pharmacare scheme modelled on the system used to control pharmaceutical costs in New Zealand should be introduced in Canada,^19-21^ making a comparison between the two countries pertinent. Nevertheless, some may question whether a comparison of health outcomes from the OECD data is appropriate because, despite the data being age-standardized, socioeconomic and other factors may differ between Canada and New Zealand. Although large differences exist between the two countries’ geographic areas and populations, over 80% of their residents live in an urban area with a similar population density (Table 2). The rates of residents with diabetes and obesity or who are regular smokers and the per capita consumption of alcohol, which are risk factors for cardiovascular disease and some cancers, are similar in the two countries. Both countries have diverse economies, with international trade being a significant component of both economies, and a similar per capita gross domestic product. In addition, both countries provide hospital and physician services without payment at the point of service, with the numbers of hospital beds and active physicians and nurses per 1,000 population in 2017 being virtually the same. Thus, comparisons are relevant and sensible.

**Table 2:** Comparison of relevant characteristics of the Canadian and New Zealand populations.

|  | Canada | New Zealand |
| --- | --- | --- |
| Residents living in an urban area<sup>22,23</sup> | 81% | 86% |
| Residents with diabetes (2017)<sup>24,25</sup> | 7% | 6% |
| Obesity rate (2017)<sup>15</sup> | 26% | 32% |
| Population aged 15+ years smoking daily (2017)<sup>26,27</sup> | 12% | 14% |
| Litres of alcohol per capita (2017)<sup>15</sup> | 8.1 | 8.8 |
| Gross domestic product per capita (US\$, 2019)<sup>28</sup> | \$46,125 | \$41,966 |
| Hospital beds per 1,000 population (2017)<sup>15</sup> | 2.6% | 2.7% |
| Active physicians per 1,000 population (2017)<sup>15</sup> | 2.7% | 3.0% |
| Nurses per 1,000 population (2017)<sup>15</sup> | 10 | 10 |

Interesting differences in health outcomes can be seen between Canada and New Zealand. In particular, mortality and hospital discharge rates in New Zealand were generally higher and the average length of stay in hospital longer for cardiovascular disorders than in Canada. This is not the result of differences in the prevalence rates of these disorders, which are similar in both countries.^29,30^

Any impact of drug access on cardiovascular mortality is likely to take many years to be seen since many cardiovascular drugs are prescribed for the treatment of conditions such as hypertension and high cholesterol levels, which are risk factors for cardiovascular mortality. A 25 to 30-year period is a reasonable length of time to make such an assessment for the ACEI and statin drugs and their impact should be observable by 2014, but the ARB drugs were introduced more recently. However, mortality from ischemic heart and cerebrovascular diseases and the rate of hospital discharges remain higher and the length of hospital stay for these health conditions is longer in New Zealand compared with Canada. The fewer ACEI and statin drugs and perhaps the ARB drugs funded and available in New Zealand may have contributed to these differences. One can argue that having more drugs in a class makes little difference, but patients are variable biologic entities so that a one-size-fits-all approach does not necessarily work. Patients may find several drugs in a class have poor effectiveness or cause adverse effects before coming to one that is effective and does not produce side effects. Finding the right drug for the right patient is necessary to promote adherence and persistency with a therapy to lead to a beneficial long-term outcome.

Unlike cardiovascular drugs, oncology drugs are not prescribed as preventative therapy but to try to alleviate an existing disorder. Despite the proliferation of cancer therapies over the past 25 years, the lack of oncology drugs in the New Zealand formulary does not appear to have negatively impacted overall cancer mortality. However, mortality from three of the four major cancers (breast, colorectal and prostate) is higher in New Zealand. Although mortality rates from lung cancer are higher in Canada in the OECD data for 1985-2014, a recent report comparing survival, mortality and incidence of seven cancers (colon, lung, oesophagus, ovary, pancreas, rectum and stomach) in seven countries, including Canada and New Zealand, between 1995 and 2014 demonstrated that the five-year survival rate for lung cancer in Canada was 40-55% higher between 2000 and 2014 than the New Zealand rate.^31^ New Zealand had lower survival rates than Canada for all the cancers in the report between 2000 and 2014, except oesophageal cancer.

## Limitations

This analysis has limitations. First and foremost, it is an associative assessment so that it is not possible to prove that lower drug listing rates lead to increased mortality and higher hospital discharge rates. Second, despite the OECD data being age-standardized making comparisons valid, other factors may have affected the health outcomes. Third, mortality rates by cause were only available to 2014 in the OECD data and, consequently, are not current. A further limitation is that listing of medications does not necessarily equate with access. Copayments, deductibles and premiums required by Canadian public drug plans,^8^ can place drugs, especially costly new ones, beyond reach. Once a drug is covered in New Zealand, this issue may be less of a problem because, in most cases, patients pay a copayment of only NZ$5 (about CAN$4.50),^32^ which is less than half the dispensing fee in most provinces before any copayments or deductibles are added. Clinical criteria for coverage of a drug also frequently restrict access.

## Conclusions

With 41.8% of new regulatory-approved medications reimbursed in 2011-2016, Canada ranks 18th out of 20 OECD countries, while New Zealand ranks 20th with just 21.8% of new medications reimbursed in the same period.^33^ Access to new drugs, including anti-hypertensives and statins, has been demonstrated in Canada and several other countries to have a beneficial impact on health outcomes that outweigh the increased cost.^34-38^ The limited access to anti-hypertensives in New Zealand is surprising because hypertension is “a worldwide problem of enormous consequence.”^39^ The PHARMAC system has contained costs in New Zealand, but it restricts or denies access to important new medicines with the potential to improve patients’ lives.

The attractiveness of a country as a priority jurisdiction in which pharmaceutical companies seek regulatory approval for their new products is based on several factors, such as the potential number of patients with the disorder for which the drug is indicated, the likelihood of obtaining a profitable price for the product and gaining drug reimbursement coverage, and the country’s overall investment climate. The ability to progress innovative treatments through regulatory, reimbursement and pricing processes in a timely manner with government officials and politicians having a comprehensive understanding of costs and system savings (not simply focusing on price) is a critical element. Canada’s complex pricing and reimbursement environment^7,8^ already places the country’s attractiveness for new innovative medications, especially costly ones, at a lower level than the United States and Europe^40^ because innovation is not seen to be valued in Canadian governments’ pricing decisions.^41^

National pharmacare and medication affordability are the current focuses of Canadian politicians, government officials and health policy analysts.^3-7^ Changes in the regulations that govern the agency that sets maximum prices for patented medicines sold in Canada will decrease the attractiveness of Canada as a jurisdiction in which global pharmaceutical companies seek regulatory approval for their new products.^3,4,42-44^ This will decrease the number of new drugs brought to Canada^45^ and, thus, reduce the number covered. A New Zealand-style national pharmacare scheme in Canada would offer public payers the opportunity to restrain drug expenditure but would likely fail to satisfy patients and healthcare providers and result in higher costs in other healthcare sectors. More research is required to understand the impact of tighter cost-containment on prescription drug coverage and health outcomes before making radical changes to the Canadian pharmaceutical environment.

## Data Availability

Data derived from public domain resources.

## Acknowledgement

The author gratefully acknowledges research funding for this work from Medicines New Zealand. The author also thanks Graeme Jarvis, Phyllida Duncan and Tanya Baker from Medicines New Zealand and Lisa Woods from Victoria University of Wellington for comments on earlier drafts of this article. Although Medicines New Zealand staff had the opportunity to review the manuscript, they had no control over the design, analysis or reporting of this work.

## Data Availability Statement

Data derived from public domain resources.

## Compliance with Ethical Standards

This article does not contain any studies with human participants or animals performed by the author.

